# Multiscale-Multistage Temporal Convolutional Network for EEG-Based Mild Cognitive Impairment Detection

**DOI:** 10.64898/2025.12.03.25341525

**Authors:** E Sathiya, Chunzhuo Wang, T. D. Rao, T. Sunil Kumar

## Abstract

Mild cognitive impairment (MCI) is an intermediate stage between normal ageing and dementia, with affected individuals at a higher risk of progressing to Alzheimer’s disease (AD). While several EEG-based MCI methods have used deep learning models, most models work at a single temporal scale, without stage-wise supervision. This limits their ability to capture the multiscale temporal structure present in EEG signals. To address this, we propose a multiscale-multistage temporal convolutional network (MS-MSTCN) for discriminating MCI from healthy controls (HC) using resting-state electroencephalography (EEG) signals. The model first processes each EEG segment through parallel dilated temporal convolution branches to capture short- and long-range patterns at multiple time scales. Three sequential stages with deep supervision then iteratively refine the class predictions at each stage, softmax probabilities are embedded and fed back into the next stage as class-conditional temporal features. We evaluated MS-MSTCN on two publicly available EEG datasets using 10-fold cross validation and compared it with two baseline models (Multiscale TCN and Multistage TCN) as well as existing EEG-based MCI methods. Across both datasets, MS-MSTCN consistently achieves superior performance and it reduces missed MCI cases and false positives (FP) in HC, which is significant for reliable early screening and clinical decision-making.

## I. Introduction

Mild cognitive impairment (MCI) is a neurodegenerative disorder, marked by cognitive decline that exceeds normal ageing, while daily functioning is largely preserved [1]. Individuals with MCI typically exhibit greater memory loss than age-matched healthy people and have a higher risk of progressing to dementia, especially Alzheimer’s disease (AD) [1]. Globally, about 55 million people are living with dementia, and it is expected to increase around 139 million by 2050 [2]. Although, there is no permanent cure for MCI, several methods are used for the diagnosis of MCI including the Mini-Mental state Examination (MSME), magnetic resonance imaging (MRI), computed tomography (CT), blood tests, positron emission tomography (PET), spinal fluid and electroencephalography (EEG) [3]. While advanced imaging and fluid biomarkers can be informative, they are costly, time-consuming, operator-dependent and less practical for repeated screening [3]. In contrast, EEG is non-invasive, portable, cost effective and it can capture neural changes associated with cognitive decline [4]. Even though many studies have performed multiclass classification involving MCI, AD and healthy controls (HC) [5,6], detecting at an early stage is crucial, as timely intervention may slow down the clinical progression and help maintain the quality of life [7,3].

Over the past decade, a wide range of machine learning (ML) and deep learning (DL) approaches have been explored for EEG-based MCI diagnosis [8,3]. The authors in [9] employed a single-channel resting-state EEG device to record 100 seconds segments from 120 participants (10 dementia, 33 MCI and 77 HC) and extracted power spectrum ratios were then fed to an support vector machine (SVM) classifier, achieving 86% accuracy (Acc) for the classification of dementia, MCI and HC. In [10] the authors used two public EEG datasets combined into 61 participants (32 HC and 29 amnestic MCI) using variational mode decomposition (VMD) based entropy features with k-nearest neighbours (KNN), they achieved up to 99.51% Acc with all 19 channels, and an ensemble KNN reached 99.81% Acc using only 11 non-dominated sorting genetic algorithm (NSGA)-selected channels. In [11] the authors used same 61-subjects, and extracted discrete wavelet transform (DWT)-based non-linear features such as band power, energy and entropy and reported 99.84% Acc with all channels, which increased to 99.97% with only 4 channels and reached 100% Acc using 5 NSGA-II-selected channels in distinguishing MCI from HC, showing that a few suitable channels will lead to superior performance in distinguishing MCI and HC. The authors in [1] used piecewise aggregate approximation (PAA) to compress the signals and extracted permutation entropy (PE) and autoregressive (AR) features which were classified using effective learning machine (ELM), SVM and KNN on a public MCI EEG database consists of 27 subjects (11 HC and 16 MCI), reaching 98.78% Acc for the detection of MCI. In [12] the authors evaluated their gated recurrent unit (GRU) model on a EEG dataset of 27 subjects (11 MCI, 16 HC), achieving 96.91% Acc, outperforming long short-term memory (LSTM), SVM and KNN classifiers. In the following study [3], the authors addressed the limitations of shallow methods by extracting deep temporal features using an LSTM model using the same 27 subjects with 5-fold cross validation for early MCI detection. The authors in [13], analysed odor-evoked multichannel EEG dataset of 35 participants (13 AD, 7 aMCI and 15 HC), combining raw EEG signals, temporal-spectral features including wavelet-based, spectral grouping, canonical correlation analysis (CCA) each were passed through an parallel convolutional neural network (CNN) branches fused with attention mechanism finally, branch embeddings were fused in a fully connected head for the classification of MCI and HC achieving an Acc of 96.17%. The authors in [14], generated spectrogram images from two EEG datasets, consisting of 27 subjects (11 HC and 16 MCI) and 109 subjects (7 MCI and 102 HC), respectively and showed that mid-to-high frequency bands are particularly informative for MCI detection, obtaining a maximum classification Acc of 99.03% with 10-fold cross validation. These studies demonstrate that various ML and DL algorithms can be effective for MCI detection. However, EEG signals are non-stationary, and in MCI, abnormalities may appear both as brief transients and as slower changes in oscillatory activity over several seconds, a model that only looks at a short window may miss slower trends, while one that only uses long windows may not capture the fine transients [1,3,12,14-15]. Therefore, capturing both short-long-range temporal patterns is important. Recent studies have successfully applied multi-scale temporal convolutional networks (MS-TCN) to brain computer interface tasks (BCI), including representation learning and motor imagery classification [16,17]. Similarly, multi-stage TCNs have been shown to progressively refine temporal predictions and reduce segmentation errors in video and gesture detection [18,19]. At the same time, temporal convolutional networks (TCNs) have been utilized for classifying other disorders [20,21]. However, to the best of our knowledge, multiscale, multistage TCN (MS-MSTCN) has not yet been explored for MCI and HC classification, where parallel dilated convolutions can provide a long receptive field and stage-wise refinement can help capture difficult patterns for distinguishing MCI from HC.

The main contributions of this paper are explained as follows:

- We proposed a MS-MSTCN model that uses multiscale dilated temporal convolutions with three-stage supervised refinement to learn both short-and long-range temporal patterns form EEG for MCI and HC classification.
- We validated the model on two publicly available EEG datasets, performed ablation using multiscale-only and multistage-only TCN variants, and showed that MS-MSTCN reduced false negatives in MCI and false positives in HC compared with these baselines.
- We also performed a state-of-the-art-methods comparison, showing that MS-MSTCN achieved a superior performance on both datasets.

This paper is organized as follows: Section II describes the EEG datasets, preprocessing pipeline. Section III details the proposed MS-MSTCN architecture together with baseline TCN variants and experimental setup. Section VI explains the evaluation results and compares the proposed model with existing EEG-based MCI detection methods then discusses limitations and future directions. Finally, Section V concludes the paper.

## II Proposed Workflow

In this study, we used two EEG datasets for MCI detection to assess the generalization of the proposed DL model. Fig.1. presents the end-to-end workflow, and the following subsections describe the datasets and acquisition used in this work.

**Fig. 1.**
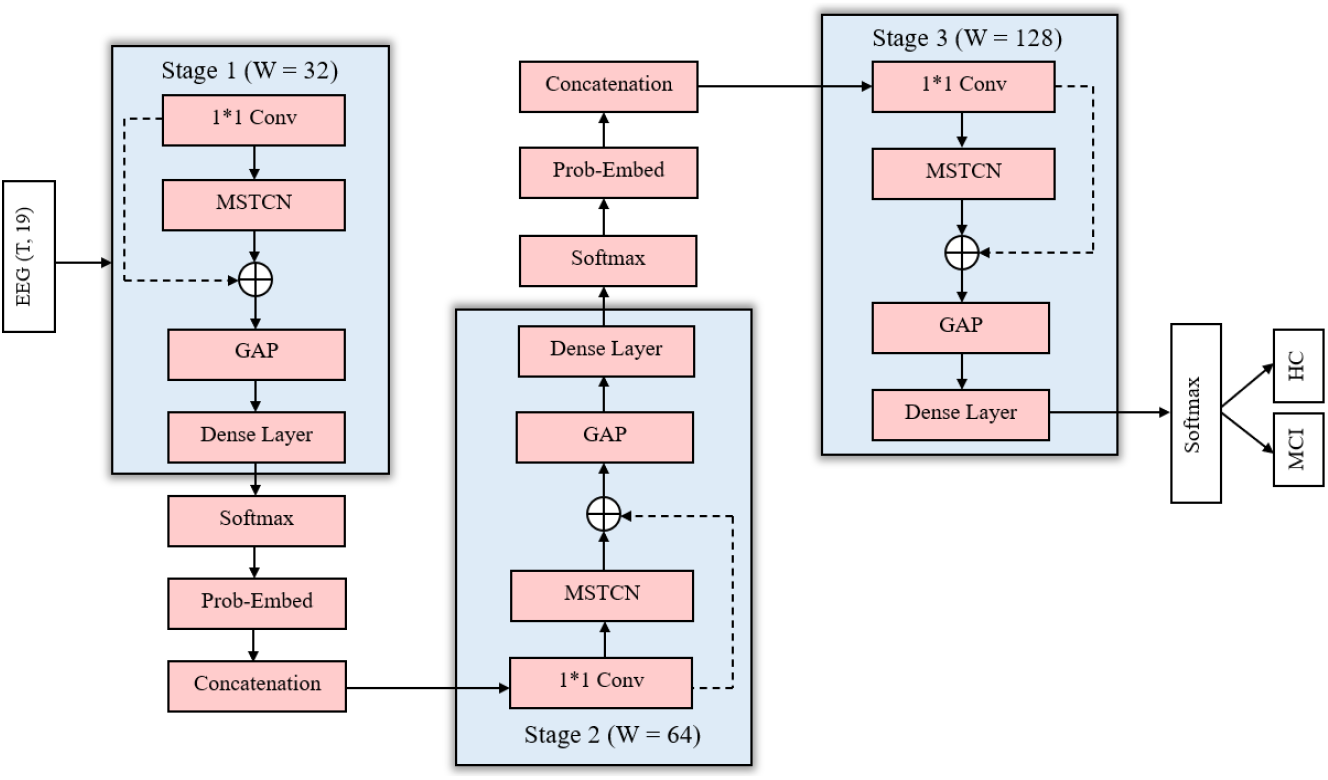
Overall architecture of the proposed MS-MSTCN for EEG-based MCI and HC classification. The network consists of three sequential stages, each containing an MSCTN block followed by global average pooling (GAP) and a dense layer. At every stage, softmax outputs are embedded and concatenated with the temporal features of the next stage, enabling multiscale feature extraction and progressive refinement before the final MCI and HC prediction. Concretely, stage s produces (i) a temporal feature vector 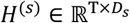, where *D*_1_ = 32, *D*_2_ = 64, and *D*_3_ = 128 and (ii) a 2D probability vector *P*^(*s*)^ ∈ ℝ^2^ over the MCI and HC classes.

### Dataset A

> We employed a publicly available dataset [22] in which EEG signals were collected at Isfahan University of medical Sciences, all participants provided written informed consent. The dataset comprises 27 older adults (age 60-77): 11 with MCI and 16 with HC. Recordings are resting-state EEG acquired on a clinical system with the sampling rate of 256 Hz. Further details about this dataset can be found in [23].

### Dataset B

> The second dataset is also publicly available, [24] introduced CAUEEG, a large EEG dataset of 1,155 patients collected at chung-Ang University (South Korea), with diagnosis assigned using standard clinical criteria and psychometric testing across 28 categories. In our study, we focus on the MCI (385 subjects) and HC (441 subjects) subsets, recorded with a sampling frequency of 200 Hz.

### Data preprocessing

EEG recordings are easily contaminated by artifacts and noise, so we applied a consistent preprocessing method to both datasets. It was performed in MNE-Python using standard 19-channel 10-20 montage. Signals were average-referenced, then a 50 Hz notch filter and a zero-phase fourth-order IIR Butterworth band-pass filter (0.5-45 Hz) were applied to remove line noise and slow drifts without introducing phase distortion. For Dataset B, we considered eyes-closed intervals identified from event logs and excluded segments with photic stimulation. For Dataset A, we used the whole resting state recording after cleaning. A window length of 1-seconds with 50% overlap was adopted following [23], where this yielded good discrimination performance on the same Isfahan dataset (Dataset A) with the same setting. For consistency, we applied the same segmentation to Dataset B as well. This resulted in 42,007 MCI and 59,846 HC windows (101,835 total) for Dataset A, and 365,365 MCI and 351,470 windows (716,835 total) for Dataset B. After windowing, each 1-seconds segment had size 256 × 19 (time samples × channels) for Dataset A and 200 × 19 for Dataset B, and each window × channel segment was z-scored (mean-zero, unit-variance) to stabilize training.

## III. Structure of the MS-MSTCN model

The proposed MS-MSTCN combines parallel TCN branches with varying kernel sizes (multiscale) and step-wise refinement across stages (multistage) as shown in Fig.1.

The architecture is composed of three sequential stages, each responsible for refining temporal representations. Internally, each stage contains an MSTCN block designed to capture temporal patterns at multiple scales using parallel temporal convolutional paths, followed by a GAP and a dense layer. Stage-wise outputs undergo deep supervision using softmax-based classification. And the predicted probabilities are embedded and fused with the original temporal features before being passed to the next stage. Thus, each stage provides both a refined temporal embedding *H*^(*s*)^ and a low-dimensional class prediction *P*^(*s*)^ that conditions the subsequent stage.

The model processes pre-processed EEG segments of shape *X* ∈ ℝ^*T*×*C*^, as defined in the Section II. In stage 1, the EEG segment first passes through a 1×1Conv1D layer and then enters the first MSTCN block. It is implemented using nine parallel branches, each configured with a unique combination of *K* = (3,5,7) and *d* = (1,2,4), forming all possible (*K, d*) pairs. Within each branch, the input undergoes two causal dilated 1D convolutions each followed by LayerNorm, GELU activation and Dropout, with a residual connection as shown in Fig. 2. The outputs of all the branches are concatenated along the feature dimension and passed through another 1×1Conv1D layer followed by LayerNorm and GELU activation. An outer residual connection then adds the original input *X* to the fused multi-scale output, forming the final stage-1 feature map 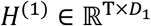 (with *D*_1_ = 32). This output is passed through a GAP layer and a softmax classifier, the GAP produces a vector 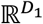, which is then mapped to the 2D probability vector *P*^(1)^ ∈ ℝ^2^. The resulting probabilities are embedded and concatenated with the temporal output *H*^(1)^ to form the input to the next stage.

**Fig. 2.**
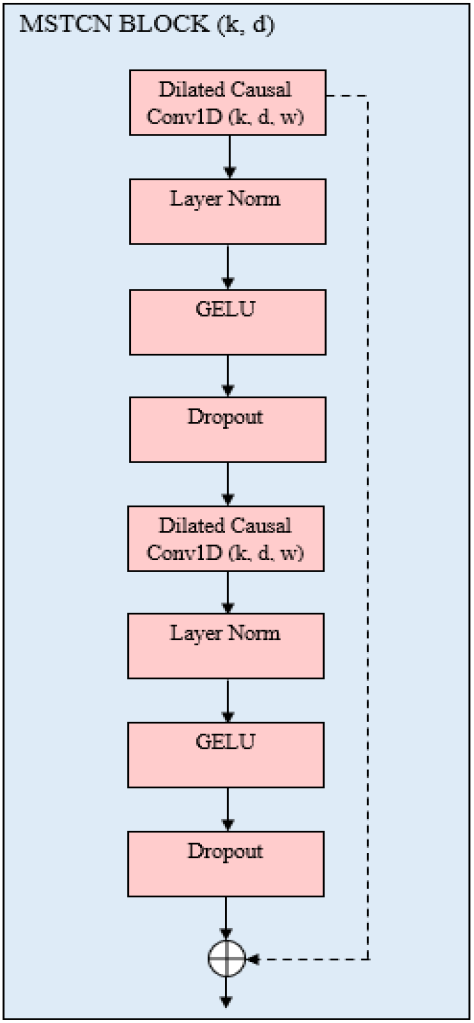
Internal structure of the MSTCN block used in each branch. The block consists of two stacked dilated causal 1D convolutional layers (kernel size *k*, dilation *d*), each followed by LayerNorm, GELU activation and Dropout, with a residual skip connection from the input to the output.

This progressive refinement continues through stage 2 and stage 3. In stage 2, the embedded probabilities *P*^(1)^ are fused with the original temporal features, and the MSTCN block outputs 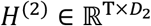 (with *D*_2_ = 64), followed again by a GAP and a 2D softmax prediction *P*^(2)^.In stage 3, the softmax probabilities from stage 2 are embedded in the same way and combined with the input *X* before feeding into the final MSTCN block, which produces 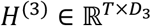 (with *D*_3_ = 128) and the final 2D softmax output *P*^(3)^. Each stage is independently supervised, allowing the network to correct earlier mispredictions and capture long-range dependencies for classifying MCI and HC subjects.

To validate the effectiveness of each architectural component, we compare MS-MSTCN with two baseline variants: multiscale TCN and a multistage TCN. The multiscale TCN retains the same parallel multibranch convolutional structure as MS-MSTCN but without any stage-wise refinement, that is, it uses a single multiscale dilated convolution block followed by a classification layer that directly outputs *P* ∈ ℝ^2^. In contrast, the multistage TCN maintains the three-stage refinement structure but replaces the multiscale modules with standard TCN blocks (using a single kernel size and dilation per layer). This allows us to isolate the effects of multiscale feature extraction and stage-wise refinement on overall performance.

### Experimental setup

For the proposed MS-MSTCN, we used three stages with one multiscale block per stage. The hidden widths per stage were set to 32, 64 and 128, respectively. Each multiscale block uses nine parallel branches given by all the combinations of kernel sizes and dilations. The dimensionality of the embedded class-conditional features was set to 8. A dropout rate of 0.10 was applied after each convolutional sub-block.

We used stratified 10-fold cross-validation at the window level and trained each model for 30 epochs with a batch size of 256. Optimization was performed with the Adam optimizer, using a learning rate of 3×10^−4^. To avoid class imbalance, the proposed MS-MSTCN was trained with class-weighted cross-entropy applied to the stage wise outputs, whereas the multistage TCN baseline keeps the three-stage refinement and conditioning structure, but replaces the multiscale modules with a standard TCN using a single fixed kernel size (k = 5) and dilations 1, 2 and 4, then uses a weighted combination of per-stage losses. The multiscale TCN baseline retains multiscale block with same kernels and dilations as MS-MSTCN with the cross-entropy loss without weights.

## IV. Results and Evaluation

To assess the performance of the proposed model, we computed mean Accuracy (Acc), Macro Precision, Macro Recall, Macro F1-score, Cohen’s *K*, Macro G-Mean across all folds, using stratified 10-fold cross-validation on the two datasets described in Section II, for MCI and HC classification. In all experiments, MS-MSTCN was compared against two baseline variants: a Multiscale TCN and a Multistage TCN.

On Dataset A, the proposed MS-MSTCN achieved an Acc of 98.86%, can be seen in Table 1, while the multiscale TCN and multistage TCN obtained 98.34% and 97.99%, respectively. Although the Acc differences are relatively small, MS-MSTCN shows clearer improvement in class-balanced metrics, with a Macro F1-score of 98.82%, Cohen’s *k* of 97.65%, and Macro G-Mean of 98.82%.

**Table. 1.** Performance of MS-MSTCN and baseline TCN variants on Dataset A and Dataset B.

| **Dataset A** |  |  |  |  |  |  |
| --- | --- | --- | --- | --- | --- | --- |
| **Model** | **Acc (%)** | **Kappa (%)** | **Macro Precision (%)** | **Macro Recall (%)** | **Macro F1-score (%)** | **Macro G-Mean (%)** |
| **MS-MSTCN** | 98.86 | 97.65 | 98.81 | 98.84 | 98.82 | 98.82 |
| **Multiscale-TCN** | 98.34 | 96.59 | 98.18 | 98.43 | 98.30 | 98.30 |
| **Multistage-TCN** | 97.99 | 95.85 | 97.84 | 98.01 | 97.93 | 97.93 |
| **Dataset B** |  |  |  |  |  |  |
| **MS-MSTCN** | 90.99 | 81.97 | 90.99 | 90.99 | 90.99 | 90.99 |
| **Multiscale-TCN** | 88.56 | 77.09 | 88.74 | 88.50 | 88.54 | 88.62 |
| **Multistage-TCN** | 86.34 | 72.64 | 86.44 | 86.29 | 86.31 | 86.37 |

On Dataset B, which includes a larger number of subjects and higher inter-subject variability, MS-MSTCN achieved an Acc of 90.99%, outperforming the multiscale TCN 88.56% and multistage TCN 86.34%. The proposed model also obtained Macro Precision and Macro Recall, Macro F1-score and Macro G-Mean of 90.99%, indicating consistent improvement over both baselines, on this larger dataset suggests that combining both multiscale temporal convolutions with stage-wise refinement improves the generalization of the model to various EEG recordings.

### Performance comparison and discussion

To further assess the effectiveness of the proposed MS-MSTCN, we compare it with several state-of-the-art methods for EEG based MCI detection.

The authors in [14] is presented in Table 2., proposed a multibranch attention-based temporal-spectral CNN for MCI detection from odor-evoked EEG, combining raw EEG signals, wavelet-based time-frequency coefficients, spectral grouping (SPG) features and CCA-derived temporal-spectral components are processed in parallel CNN branches for MCI and HC classification. In [25] the authors employed a MCI diagnostic model that combines iterative amplitude adjusted Fourier transform (IAAFT)-based data augmentation, sample entropy features, and a BiLSTM classifier on small resting-state EEG datasets, showing that it can improve the model performance but at the cost of extra preprocessing and training time. The authors in [3,13] proposed a deep learning framework for MCI detection in which denoised, segmented EEG is classified using LSTM and GRU models, with GRU giving the best outcome. In contrast, our MS-MSTCN focuses on multiscale temporal convolutions with multistage refinement, learning richer temporal patterns without relying on recurrent units. Similarly, in [26] the authors employed a dual attention assisted compact convolutional network with stacked Bi-LSTM (DCCN-SBiL), which combines several optimization steps for MCI detection. It is worth noting that [3,13] used the same Dataset A as in our work, and achieved lower performance compared with our MS-MSTCN model. In addition, unlike these studies, which are restricted to a single small dataset, we also evaluated MS-MSTCN on a larger EEG dataset, providing a practical alternative to existing methods while reducing clinically important errors such as missed MCI cases.

**Table 2.** Comparison of the proposed MS-MSTCN with recent state-of-the-art EEG-based MCI detection models.

| **Ref** | **Dataset** | **Model** | **Acc (%)** |
| --- | --- | --- | --- |
| [14] | 13 AD, 7 MCI and 15 HC | Multibranch attention-based temporal-spectral CNN | 96.17 |
| [25] | 10 MCI, 10 HC | BiLSTM | 97.2 |
| [3] | 11 MCI, 16 HC | LSTM | 96.41 |
| [13] | 11 MCI, 16 HC | GRU | 95.51 |
| [26] | 15 HC, 7 MCI and 13 AD | DCCN-SBiL | 95.3 |
| **Proposed** | **11 MCI, 16 HC and 459 HC, 417 MCI** | **MS-MSTCN** | **98.86 and 90.99** |

Note : BiLSTM: Bidirectional LSTM; DCCN-SBiL : dual attention assisted compact convolutional network.

Although MS-MSTCN achieves strong classification performance, its primary limitation is increased time complexity due to the use of multiscale convolutional branches and the multistage refinement blocks, which may affect deployment in real-time or low resource environments. As a future work, we aim to integrate explainable AI (XAI) techniques to improve interpretability of the model’s decisions, thereby supporting clinical validation and enhance trust in automated MCI diagnosis.

## V. Conclusion

In this work, we proposed an MS-MSTCN model for EEG-based MCI detection from resting-state signals, combining multiscale dilated temporal convolutions with multistage refinement to better capture temporal patterns. The model was evaluated on two EEG datasets and consistently outperformed multiscale TCN and multistage TCN baselines. Ablation and confusion matrix analyses showed that using both multiscale and multistage components together reduces FN in MCI and FP in HC, with the improvements being more evident on the larger dataset, indicating better generalization.

## Data Availability

All data analyzed in this study are from previously collected EEG datasets. The Isfahan MCI EEG dataset is publicly available at https://misp.mui.ac.ir/en/eeg-data-0. The CAUEEG dataset is available to qualified academic researchers upon reasonable request to the original authors, access instructions are provided at https://github.com/ipis-mjkim/caueeg-dataset. No new human data were collected by the authors for this study.

https://github.com/ipis-mjkim/caueeg-dataset

## Notes

### Competing Interest Statement

The authors have declared no competing interest.

### Funding Statement

This study did not receive any funding

### Author Declarations

Ethics committee/IRB of Isfahan University of Medical Sciences, Isfahan, Iran, gave ethical approval for the original EEG recordings used to create the Isfahan MCI EEG dataset. The Institutional Review Board of Chung-Ang University Hospital, Chung-Ang University, Seoul, Republic of Korea, gave ethical approval for the original EEG recordings in the CAUEEG dataset. This work involves only secondary analysis of these fully anonymized, previously collected datasets, no new human data were collected by the authors and no identifiable information was available, so no additional ethics approval or consent were required for this study.

